# Capturing cultural and linguistic diversity in child health research in Australia

**DOI:** 10.1101/2020.10.25.20219329

**Authors:** Razlyn Abdul Rahim, Rhiannon Pilkington, Katina D’Onise, John Lynch

**Author notes:** Corresponding author: (RAR).

## Abstract

The concept of Culturally and Linguistically Diverse (CALD) populations is unique to Australia. It was introduced in 1996 and is intended to refer to ethno-cultural groups that are neither Aboriginal or Torres Strait Islander nor considered from mainstream English-speaking Anglo-Celtic backgrounds. CALD children have been identified as a priority population by the Australian government because they may experience inequities in health outcomes compared to Anglo-Australian children. Inequities in the health and wellbeing of CALD children are driven by myriad processes including racial discrimination, socioeconomic disadvantage, and limited access to health services. But who are CALD children? Despite the availability of statistical standards for data collection on CALD characteristics such as country of birth and language spoken, the concept itself lacks an official operational definition. Applying definitions specified by various organisations to data from the 2016 Australian Census, the estimated proportion of CALD children ranged from 11% to 44% of Australian children aged 0 to 17 years. There are few published studies on CALD children in Australian child health research, with most studies focused on refugees. There is no consensus on how CALD is defined in child health research in Australia. We propose several considerations in the use of the CALD concept in child health research. This includes adhering to the Australian Bureau of Statistics standards on Cultural and Linguistic Diversity, use of multiple indicators to identify CALD, and acknowledging the significant heterogeneity of CALD communities which may contribute to observed differences in health. If we are to advance health and well-being equity for CALD children, we need a more carefully considered and consistent approach to understanding which children are CALD.

## Introduction

Twenty-first century Australia is claimed to be one of the most multicultural societies in the world. However, diversity brought about through waves of immigration has not always been celebrated. Australian history can be characterised by mainstream Anglo population’s domination over groups perceived as ‘others’ [1]. This domination was achieved through discriminatory policies and practices beginning with colonial governments, and was justified by racist ideology. These ethno-cultural ‘others’ refer to ethnic or cultural groups perceived as different from mainstream Anglo-Celtic Australians. One of the more recent manifestations of this desire to describe the ‘other’ is through using the concept of Culturally and Linguistically Diverse (CALD) populations. CALD is a concept unique to Australia. CALD refers to Australians who are not of the mainstream English-speaking Anglo-Celtic group and are not Aboriginal or Torres Strait Islander. Although Aboriginal and Torres Strait Islander people are diverse in language and customs, they occupy a unique position as First Australians [2], and therefore should be considered distinct from the CALD population.

CALD children, including those of refugee and asylum seeker background, are identified as a priority population in the *National Action Plan for the Health of Children and Young People 2020-2030* by the Australian government [3]. This is because the health and wellbeing experience of this group can differ from mainstream experiences. Inequities in child health and wellbeing are driven by fundamental social, political and economic factors such as racial discrimination, socioeconomic disadvantage and limited access to health services [4]. We cannot begin to understand the characteristics and extent of health inequities experienced by CALD children if we cannot consistently identify and enumerate them in data collection for health system and research purposes. Identification and enumeration of CALD children is required to inform the development of policies and appropriate service delivery to better meet their health and wellbeing needs. However, difficulties in identifying and enumerating CALD complicates the endeavour to better understand the health experiences of CALD children.

In this paper, we first trace the evolution of how the construct of the ethno-cultural ‘other’ has been reflected in conceptualizing and collecting demographic information to characterise CALD. We briefly compare methods of enumerating population ethno-cultural information in countries of similar cultural-political background to Australia. We acknowledge that while the historical description on ethno-cultural relations may be distressing, it is important and relevant to the topic. We use data from the Australian Bureau of Statistics (ABS) 2016 National Census of Population and Housing and the Australian Census and Migrants Integrated Dataset 2016 to estimate the proportion of children in Australia who could be considered CALD. Finally, we explore how the CALD population has been represented in the research on child health in Australia.

### A brief history of the construct of ‘otherness’

First described by the German philosopher Georg Wilhelm Friedrich Hegel, the ‘other’ or ‘otherness’ is a construct used to create distinction from the ‘self’ [5]. According to Hegel, the essence of self-consciousness is ‘I’ and the ‘other’ is perceived as an ‘unessential, negatively characterised object’ (p. 113)[5]. French philosopher and feminist Simone de Beauvoir extended Hegel’s idea in her seminal work, ‘The Second Sex’. de Beauvoir stated ‘…alterity is the fundamental category of human thought’ (p.26) and that in defining itself, all groups set up the other as its opposite, and views the other with hostility [1]. Examples of this opposing dichotomy include men/women, colonial/indigenous, natives/foreigners [1]. At its most apparent, the distinction is based on sensory evidence of differences in appearance or behaviours, which is perceived as foreign from the ‘self’ and therefore labelled ‘other’. There are many potential dimensions of the ‘other’ e.g., sex, sexual orientation, disability, and socioeconomic position [6]. The dimension under consideration here is ethno-cultural ‘other’ which relates to ethnic groups different to the mainstream ‘self’. ‘Otherness’ is an inherently hierarchical construct whereby the ‘self’, with its known and familiar characteristics, is benchmarked favourably over the ‘other’ whose features are unknown, unfamiliar and therefore assumed inferior in comparison [1, 5, 7, 8]. The implicit tendency to position the ‘self’ as superior and relegating the ‘other’ to inferior has occurred in most societies [9]. Ancient Egyptians referred to themselves as ‘the people’ and ancient Greeks refer to ‘others’ from geographically distinct civilisations as ‘Barbarians’ [8, 10]. This notion of othering, known as ethnocentricity, is based on the premise that ‘others’ are not socialised into the language and culture of the ‘self’, but could be accepted as adopted society members if they assimilated [9]. These ancient historical ideas of ethnocentricism differ from more modern racial ideology, because racist ideologies focus on intrinsic, fixed characteristics which deem that the inferior ‘other’ could never be part of the ‘self’ [9].

‘Otherness’ is socially constructed based on group identity [6]. It is a complex and nebulous construct because human diversity is a product of geographical, historical, sociological and anthropological influences that is unlikely to be captured with any single indicator. Various contemporary indicators used to describe the ethnocultural ‘other’ have been imperfect, with classification categories that have tended to conflate separate entities such as race, ethnicity, nationality or historical and geographic origins [11-14].

### Race

The concept of race as an indicator of ethno-cultural ‘otherness’ is a relatively modern development within the last 400 years [9]. The word race originated from the Spanish word *raza* which means breed or stock [15]. ‘Race’ is rooted in essentialism [16], and refers to so-called fixed, inherent biological or phenotypical characteristics shared by a group of people, and is defined and imposed by others [11, 12, 17]. Now widely discredited as a biologically meaningful construct, ‘race’ categorised humans into what were believed to be distinct and naturally hierarchical clusters [15, 18-20]. Work to classify humans into distinct groups became part of scientific theory during the Enlightenment [21]. Carl Linnaeus, a Swedish botanist, zoologist and physician, first introduced a Human Taxonomy in 1735 where humans were coded as white Europeans, red Americans, dark Asians (revised as yellow) and black Africans [20, 22]. In 1795, Johann Friedrich Blumenbach described five varieties of humans - Caucasians, Mongolians, Ethiopians, Americans and Malays [23]. Blumenbach emphasised geographical gradation between populations and that the five varieties of humans belong to one species [23]; however according to Bhopal this was displaced in favour of the notion there are multiple species of humans, an idea widely held in the 18^th^ century [24].

The ideology of races as hierarchical was consolidated as a result of European interaction with other populations through conquest, colonialism and the slave trade in Africa, Asia and the Americas [9, 15, 16, 19, 25, 26]. During the 17^th^ century it was believed that ‘negroes and savages’ were half animals [24] and intrinsically inferior to white people in intelligence and morality [19, 22]. Arthur de Gobineau, a French diplomat, writer and ethnologist, published an ‘Essay on the Inequality of Human Races’ in 1853 which hierarchically described three primary races of man as ‘white’, ‘yellow’ and then ‘black’ [27]. Gobineau, whose work influenced Nazism, argued the inferior ‘black’ and ‘yellow’ races did not have the intellect to establish civilisations compared to the ‘…immense superiority of the white peoples…’ (p.207)[27]. This biologic determinism drove social relations that justified subordination of non-White people, allowing for the oppression of such groups through slavery and colonialism [16, 18, 19, 22, 25]. For example in the United States, people of European descent were full citizens whilst those of African descent were slaves [16]. Heavy economic reliance on slave labour reinforced essentialist beliefs of the lesser status of African people [16]. Similarly, the construction of Orientalism in the 18^th^ century provided a system to understand, create knowledge about, and govern the ‘Orient’ by Franco-British colonial rulers [28]. Steeped in the dogma ‘Orientals’ were incapable of self-government, colonial rule was justified to benefit the ‘Orientals’ and ‘…also for the sake of Europe at large’ (p.33)[28].

Scientists and physicians reinforced racial ideology by conducting studies and writings to ‘prove’ the inferiority of non-whites for example through comparing the skull shapes and sizes of different groups and conducting experiments on black slaves [26, 29]. These pseudoscientific theories dominated European thinking in the 19^th^ century to consolidate the European and ‘others’ dichotomy [9]. Scientific racism continued up to the mid-20^th^ century with the eugenics movement – meaning ‘well born’. Eugenics was conceived in the late 19^th^ century by British scientist Sir Francis Galton [30]. These efforts to provide a scientific basis to racial inferiority helped underpin well-known social policies and crimes against humanity such as the Jewish Holocaust. In the wake of the atrocities of World War II, a consensus was reached that the view of race as fixed and hierarchical was socially created with no biological basis [8].

The advent of genomic science in the late 20^th^ century provided further evidence disputing scientific racism. Genetics offers no biological evidence to support natural hierarchies between various population groups [15]. Genetic variation observed between population groups supports the hypothesis that humans share a common ancestor that appeared in Africa more than 150,000 to 200,000 years ago [31, 32]. A landmark study published in *Science* in 2002 demonstrated that genetic variation within a population accounted for 93-95% of genetic diversity; and variation between major population groups makes up only 3-5% [33]. There is more genetic diversity observed in a group of people from the same geographic region than between population groups of major geographic regions of Africa, Eurasia, East Asia, Oceania and America [33]. Six main genetic clusters - subgroups with ‘distinctive allele frequencies’ (p. 2382) - were identified corresponding to major geographic regions, suggesting migration patterns out of Africa [33]. Genomics confirmed Blumenbach’s description of geographic gradation or ‘clinal’ patterns in human populations. That is, the genetic profile of individuals demonstrate partial membership in multiple clusters thus nullifying the notion of race as distinct groups [15, 32-34]. Skin colour, the major phenotypic component used to discriminate groups, is more varied between geographic groups than within. Differences in skin colour measured by skin reflectance in samples from five major geographic regions, (i.e. Sub-Saharan Africa, Europe, Central/East Asia, Australasia, the Americas) varied by 88% between these regions compared to 9% variance within regional local populations [35]. This is likely explained by selective pressure where those with ancestral origins in locations proximal to the equator have darker skin tones as a protective mechanism against the effects of more intense sun exposure, compared to those distal to the equator [15, 35].

Contemporary research provides no biological evidence to support different races being natural categories as posited by past pseudoscientific evidence. Thus, it is widely accepted that race, with its unchanging and biologically hierarchical basis, is a socially engineered construct intended to justify European political and economic hegemony in the last 400 years.

### Ethnicity

As racial ideology became increasingly discredited, from the 1970s the concept of ‘ethnicity’ was advocated as a way to characterise people [22, 31, 36, 37]. The term ‘ethnicity’ was derived from the Greek term *ethnos* or nation, and is a complex, multifaceted construct [31]. Moving away from seemingly innate, biological characteristics, ethnicity expanded the components that define groups to include geography, ancestral origin and sociocultural elements such as language, religion, and customs [12, 17, 31, 38]. Core to the concept is identifying common origins or social background and shared distinct cultural elements, language or religion [39]. In contrast to race which was assumed immutable, ethnicity is dynamic and may change depending on individual self-perceptions influenced by generational change and socio-political context [12, 13, 17, 31, 40-42]. The complexity and contextual nature of this construct makes ethnicity difficult to measure or assess in many populations [39].

### Ethnicity recording in Australia

In the first half of the 1900s, enumeration in Australia occurred in the form of racial classification [17]. ‘Race’ or ‘racial origin’ was recorded from the first Australian census in 1911, through to 1976. Recording was initially driven by the White Australia policy (1901 – 1966) to enumerate people of non-European and partly non-European descent and distinguishing them from the White, European or British populations [12, 43, 44]. This highly discriminatory policy was embedded in racist doctrines upheld by the colonial government which also declared Australia as *terra nullius*, disregarding the rights, laws and customs of Aboriginal and Torres Strait Islander peoples as first inhabitants of the land [45]. Data collection methods have predominantly moved away from racial classifications but in some jurisdictions such as Western Australia and, until recently, South Australia, ‘race’ was still recorded as a category in routinely collected perinatal statistics [46, 47].

By the 1970’s the White Australia policy was repealed and replaced by multiculturalism with underlying principles of access, equity and participation [48]. This was a watershed moment. For the first time in Australia, a policy framework was established to meet the needs of immigrants of diverse backgrounds [49]. Accompanying this change, data items like parental place of birth, language spoken, ancestry, and English proficiency were introduced in the Census which enabled enumeration of diverse groups to inform anti-discrimination policies [43]. The 1986 Population Census Ethnicity Committee endorsed the

UK Law Lord’s definition of ethnicity which formed the basis of the Australian Standard Classification of Cultural and Ethnic Groups (ASCCEG)[40]. Characteristics that could be considered in this definition of ethnicity were a distinct shared history, cultural tradition, geographic origin, language, literature, religion, minority status and being racially visible [40]. The ASCCEG forms the classification for coding of ancestries identified by Census respondents. When used in combination with other information such as country of birth and parental country of birth, ancestry information provides an indication of the respondents’ ethnicity [50].

Despite the policy of multiculturalism, Australia’s national identity has remained focussed on British heritage. A concept used to describe non-British people in Australia in the 1980s and 1990s was ‘non-English speaking background’ (NESB)[51]. Limited information is available on how this indicator was collected and determined. Although not widely used now, remnants can still be found for example the Australian Public Service Commission website in 2017 describes NESB to include characteristics about first language spoken, country of birth and language spoken by the person’s parents [52]. NESB was criticised as it did not distinguish between disadvantaged populations, or various cultural and linguistic groups in Australia [53].

In 1996, The Ministerial Council of Immigration and Multicultural Affairs recommended the term NESB be replaced by CALD [53, 54]. Following extensive consultation, the ABS developed ‘Standards for Statistics on Cultural and Language Diversity’ which recognises the multidimensional aspects of ethnicity. These standards include a minimum set of four core indicators – country of birth, main language other than English spoken at home, proficiency in spoken English, and whether the person is Aboriginal or

Torres Strait Islander (as stated, Aboriginal and Torres Strait Islander peoples are not considered CALD peoples). Additional variables that may be included in this standard are ancestry based on geographic origins or common social and cultural characteristics shared by a group of people [55], country of birth of father and mother, first language spoken, languages spoken at home, main language spoken at home, religious affiliation, and year of arrival to Australia [53, 56].

Despite declaring this as a ‘standard’, the ABS provides no clear definition of the concept of CALD outside of four core and eight additional indicators. There is no detailed advice on how these variables should be used or combined as this is apparently contingent on the specific objectives of individual organisations [56]. However, the ABS stipulates that no single indicator would accurately synthesise different cultural and linguistic characteristics of an individual or a group of people. (p.2) [56]. In the absence of a single definition, organisations that have reported on CALD people have specified their own. These definitions differ but revolve around English language use. A list of definitions by organisations is available in Table S1.

### Ethnicity recording in the UK, Ireland, USA, Canada and New Zealand

By way of contrast, we compare Australia’s census data collection with other countries of predominant Anglo-Celtic populations namely the UK, Ireland, New Zealand, USA and Canada [57]. All countries permit self-declaration of more than one option to enable census ethnicity data collection in a sensitive way as stipulated by the United Nations [38]. All five countries have a direct question on race or ethnicity with response categories dominated by phenotypic features, geographic region of origin or cultural similarities. Australia’s question on ancestry is akin to ethnicity, but differs from the others by not using phenotype indicators such as White and Black. In the UK and Ireland ethnic categories are broadly classified by a combination of skin colour and geographic origins e.g. White, Black/African/Caribbean, Asian, Mixed and Other [58-60]. The USA has a question on race with categories White, Black/African American, Asian, American Indian/Alaskan Native, Native Hawaiian or Other Pacific Islander and some other race [61, 62]. This question is supplemented by another on Latino, Hispanic or Spanish origin [62]. Canada has two questions on ethnicity. One asks about ‘population group’ identification listing categories such as White, Black, West Asian and Latin American [63]. The other question on ethnicity is akin to the question on ancestry in the Australian census where respondents nominate one or more ethnic group eg. Greek, Vietnamese [63, 64]. New Zealand collects information on ethnic origin broadly categorised into geographic locations or cultural similarities eg. European, Maori, Asian [65].

Except for the USA, all countries include other indicators of ethno-cultural diversity in their census. New Zealand, Ireland, the UK and Canada, like Australia, collect information on person’s country of birth and languages spoken other than English (and French in Canada). Like Australia, the UK, Ireland and Canada also collect information on English proficiency. Religious affiliation is collected by New Zealand, UK and Ireland. Although Australia collects a suite of information to characterise cultural and language diversity, Canada’s data collection standards for population diversity are the most comprehensive as it also includes immigration details for permanent residents and various identification for Aboriginal and First Nation communities [66]. The census collections within these countries recognise the complexity of characterising cultural and language diversity, and its’ evolving nature in the context of migration and acculturation. As such there is increasing use of a composite of indicators to canvas ethno-cultural diversity.

The concept of CALD is unique to Australia and is intended to move away from the stigma attached to using phenotypic features to classify people. As such CALD may not be fit for use as a measure of discrimination based on appearance. In contrast, the Canadian concept of ‘visible minority’ may better encapsulate the experience of racism. The *Employment Equity* Act in Canada defines visible minorities as ‘persons, other than Aboriginal persons, who are non-Caucasian in race or non-white in colour’ and specifically consist of South Asian, Chinese, Black, Filipino, Latin American, Arab, Southeast Asian, West Asian, Korean and Japanese [67]. ‘Visible minority’ has been criticised for placing the groups it refers to in the margin compared to the dominant group, and assumes that groups in the category have a homogeneous experience of discrimination [68], highlighting the dynamic nature of ethno-cultural identification that may vary according to the sociocultural context.

The approach to enumerate the ethno-cultural ‘other’ in Australia has transitioned from one driven by racist ideology to that which aims to highlight diversity and promote inclusivity. Although there are statistical ‘standards’ for collecting information on elements of cultural and language diversity, there is no clear guidance on how these standards are applied to determine CALD identification.

### Methods

We undertook three empirical pieces of work. First, we explored how different definitions of CALD affect the size of the CALD population of Australian children aged 0 to 17 years using the 2016 ABS Census of Population and Housing. Second, we examined another dimension of the CALD construct, by estimating the number of migrant children and youth aged 0 to 19 years according to permanent visa category. And third, we reviewed the Australian child health literature to explore how CALD has been used in research.

The 2016 Census includes all people in Australia including visitors and excluded foreign diplomats and their families, and Australian residents overseas on Census night [69]. Additionally, we used the Australian Census and Migrants Integrated Dataset (ACMID) 2016 to offer a different view of children who are potentially CALD as a contrast to CALD definitions that can be applied to Census data. ACMID 2016 includes Australian Census 2016 data linked to Permanent Migrant Settlement Data from the Department of Home Affairs on migrants granted a permanent visa between 1 January 2000 and 9 August 2016 [70].

Descriptive analysis was conducted using TableBuilder Pro publicly available following application to the ABS. The 2016 Census data counting persons and place of usual residence was used for ages 0 to 17 years. Cultural and language diversity variables country of birth, Indigenous identification, language spoken at home and spoken English proficiency were used to estimate the CALD population using different definitions. ACMID 2016 was used to estimate the number of individuals based on permanent visa categories. Available data does not permit customised age groups. Hence, we present the distribution of permanent visa categories for migrant children and youth aged 0 to 19 years as proportions of migrant children, and of all children in Australia. The project was exempted from formal ethical review by the University of Adelaide Human Research Ethics Committee due to the use of publicly available non-identifiable data held by the ABS which involved negligible risk to participants.

## Results

### CALD children in the Australian 2016 Census

Table 1 presents the estimated size of CALD and non-CALD child populations in Australia according to various CALD definitions. In 2016 there were 5,208,022 children aged 0 to 17 years in Australia. Based on five definitions of CALD outlined in Table 1, the estimated proportion of CALD children in Australia in 2016 ranged from 11% to 44%. A definition based only on country of birth yielded an estimate of 11%. Definitions based only on country of birth were utilised by the AIHW and FECCA in reports on the aged care needs of older CALD Australians. Applying this definition resulted in a potential undercount of children born in Australia who may identify as CALD in other ways such as by speaking a language other than English at home.

**Table 1.** Number and percentage of children (0-17 years) according to different definitions of CALD using 2016 Australian Census of Population and Housing data (N = 5,208,022)

| Definitions of CALD | CALD children<br>n (row %) | Non-CALD children<br>n (row %) |
| --- | --- | --- |
| i) Country of birth is NOT Australia and its external territories, New Zealand, the UK, Ireland, the USA, Canada or South Africa | 574,011 (11%) | 4,634,011 (89%) |
| ii) People born in Australia whose main or preferred language spoken is not English | 1,429,959 (27%) | 3,778,063 (73%) |
| iii) Combining criteria i and ii above | 2,003,970 (38%) | 3,204,052 (62%) |
| * Definition used by the Australian Institute of Health and Welfare (AIHW) |  |  |
| Born overseas in a non-English speaking country (ie. Not born in Australia, NZ, UK, Ireland, USA and Canada<sup>^</sup>) | 592,718 (11%) | 4,615,304 (89%) |
| * Definition used by Federation of Ethnic Communities Council of Australia (FECCA) |  |  |
| Australia's non-Aboriginal and Torres Strait Islander cultural groups who are not from an English-speaking, Anglo-Celtic<sup>□</sup> background | 2,308,478 (44%) | 2,899,544 (56%) |
| * Definition used by Department of Social Services (Commonwealth Government), and Office of Multicultural Interests (Western Australia) |  |  |
| i) Non-Aboriginal or Torres Strait Islander<br>ii) Country of birth – any country<br>iii) Language spoken at home is not English<br>iv) Any level of English language proficiency<br>* Definition recommended using 'core variables' of CALD standards by the Australian Bureau of Statistics | 1,262,869 (24%) | 3,945,153 (76%) |

<sup>^</sup> FECCA does not specify which countries are non-English speaking therefore this has been interpreted to exclude South Africa where the Anglo Celtic community constitutes a minority of the population
<sup>□</sup> includes categories of ancestries: Australian, New Zealander, British, Irish, American and Canadian.
Respondents that nominated a single ancestry or dual ancestry comprising of a combination of these categories were considered Anglo-Celtic

Use of multiple indicators increases the proportion of children identified as CALD. When the CALD definition by the Department of Social Services and Office of Multicultural Interests was applied the proportion of CALD children in Australia was 44%. Here CALD children were those non-Aboriginal or Torres Strait Islander, spoke English and another language and whose ancestries excluded Australian, New Zealander, British, Irish, American or Canadian, either as a single ancestry or as combinations of these. We also applied a combination of the ABS CALD standard four core variables which resulted in CALD children comprising 24% of children aged 0 to 17 years in Australia in 2016. As the ABS does not specify how standards on country of birth and English language proficiency identified CALD, we determined CALD children as non-Aboriginal or Torres Strait Islander, spoke a language other than English at home, was born in any country and reported any level of English proficiency.

### Migrant children in the ACMID 2016

Over two thirds (68%) of Australian migrant children and youths granted visas between the years 2000 and 2016 arrived under the skilled migration program. This group comprised 5% of all those aged 0 to 19 years in the 2016 Census. Refugees resettled under the Australian Humanitarian Program accounted for 14% of all permanent residents arriving between 2000 and 2016; representing 1% of the total number of individuals aged 0 to 19 years in Australia. Although a small proportion, refugee children and youth are highlighted as a priority subgroup within the CALD population as they may experience higher levels of health and social disadvantage compared to skilled migrants due to their pre-migration circumstances. See Table S2 for the distribution of migrant children and youth by permanent visa categories.

### Health and wellbeing research on CALD children in Australia

If CALD children are a ‘priority’ population group [3] it is imperative they are well identified in health research and administrative datasets to inform improved, equitable service delivery. ‘Priority’ populations “… are especially prone to health inequity as a result of social, geographical and other determinants” (p.8); and for CALD children these may include cultural barriers to accessing health services [3]. Additionally, traumatic experiences that occur pre-settlement may have significant impact on the physical and mental health of refugee children [3].

A systematic literature search on *PubMed and Web of Science* using the terms ‘CALD’ or ‘culturally and linguistically diverse’ children, adolescent or youth health or wellbeing, in Australia from 1996 to 2019 produced 46 publications. Although the term CALD was introduced in 1996, our search yielded only two peer reviewed publications prior to 2010 that used this term. The frequency of publications using CALD increased after 2010. Nine of the 33 publications in *Web of Science* using CALD were published in 2019, compared to two in 2010 and three in 2011. To explore how ethno-cultural groups were described, an additional two searches were undertaken using the terms ‘ethnic minority’ and ‘migrant’. These searches respectively returned 22 and 128 articles. However, all these search strategies exclude publications on refugee children who are of CALD background. A search on the term ‘refugee’ on the two databases produced 176 publications focussed on the health and wellbeing of refugee children, adolescents and youths in Australia.

Of the 372 peer reviewed publications on the health and wellbeing of children from CALD backgrounds in Australia, nearly half were focussed on refugees and asylum seekers, particularly new arrival refugee children. The visibility of this group and consequent research focus may be explained by contentious debates on humanitarian intake quotas, offshore processing and detention of asylum seekers and the health and social needs of refugees. It has been suggested research concentrates on new arrivals as they are perceived to be ‘most’ different from the mainstream Anglo-Celtic Australian population [51]. Less visible are CALD children who are not refugees. It is also possible the ‘healthy migrant effect’ [71] may result in migrant children having fewer encounters with the Australian health system thereby contributing to the underrepresentation of CALD children in health research.

### How was CALD defined in child health and wellbeing research?

There is no consensus on a definition of CALD in the 46 publications that used this term. Publications mostly utilised ABS standards to identify CALD. ‘Language spoken other than English’ was the most commonly used indicator to define CALD (11 of the 46 publications) and 15 used language variously combined with country of birth, English proficiency, length of stay in Australia, religion and visa type (distinguishing migrant from refugee). Six articles solely used country of birth (i.e. born overseas or born in a non-English speaking country) to determine CALD. Three articles used ‘cultural group’ or ‘background’ which, along with visa type, are not consistent with ABS CALD standards. Two studies directly asked study participants if they identified as CALD. A publications list by how CALD was defined is available in Table S3.

A review of the collection of indicators of ethnicity and cultural diversity used in the Australian health system reveals variation in data collection [72]. National data dictionaries for health, community services and housing assistance sectors endorse cultural and language diversity information standards as stipulated by the AIHW [72]. However, national minimum dataset specifications, national health data collection structures and national surveys do not consistently apply the minimum core of the ABS Standards on Cultural and Language Diversity in collecting data on ethno-cultural characteristics [72]. In the health sector, there is emphasis on country of birth and language spoken; and a preference for information that supports service provision such as need for interpreter and preferred language [13, 72].

### Health and wellbeing issues studied

Studies conducted in Australia on CALD, ethnic minority or migrant children and parents health and well-being published since 2010 (n=65) fall under various themes. Common themes include patterns of chronic disease risk factors such as obesity, diet and physical activity (22% of publications), mental health and wellbeing (14%), access/utilisation of health and community services (14%). Studies on refugee and asylum seeker children and adolescents in Australia published between 2010 and 2019 (n=139) predominantly investigated themes such as mental health and wellbeing (27% of publications), physical health issues of various non-communicable and communicable diseases (17%), access/utilisation of services and service delivery models (13%), and wellbeing associated with resettlement (14%). A list of publications by theme is available in Tables S4 and S5.

A large number of these studies were descriptive pieces focussed on health condition patterns with no comparison made to the general Australian population. However, studies that compared CALD with non-CALD children demonstrated the higher occurrence of health and wellbeing concerns in the CALD group. For example, CALD children are more likely than their non-CALD counterparts to be overweight or obese [73-75], to have lower object control skill (an indicator for gross motor development) [76], less physically active [77] and were more likely to experience emotional difficulties and peer problems [78]. One study found CALD children who presented to hospital were more likely to leave prior to clinician authorised discharge compared to non-CALD children [79].

## Discussion

CALD, a concept unique to Australia, is intended to canvas the ethno-cultural diversity of the population. However, despite its official position in Australian population enumeration methods, the usefulness of this concept in describing ethno-cultural characteristics remains unclear. CALD in itself is partial, as the complexities of human groupings cannot be exhaustively captured nor clearly categorised by statistical criteria. We have demonstrated the difficulties of operationalising this complex concept based on recommended standards. Depending on the definition used the estimated proportion of CALD children ranged from 11% to 44% of the total population of Australian children aged 0 to 17 years. Using four core ABS indicators showed 24% of children aged 0-17 in Australia were CALD.

Another view of CALD is highlighted by visa categories where humanitarian entrants accounted for 14% of the proportion of migrant children and youth, and 1% of all Australians aged 0 to 19 years. Similarly there is no consensus in defining CALD in child health research. The few studies that compared outcomes between CALD and non-CALD children demonstrated that CALD children experience various forms of health disadvantage. How CALD is enumerated effects estimates of the population which, in turn, may bias estimates quantifying the health issue of interest in CALD communities. Although there is no single correct way in defining CALD, consistency in enumerating this population contributes to improving the quality of evidence generated that inform health policies and services developed to meet the needs of this priority population.

Here we propose several points for consideration for the use of the concept of CALD in health research in Australia. First, researchers should utilise the ABS standards on CALD as it provides a consistent method of collecting and statistically categorising information. Studies examining CALD populations using specially designed datasets should ensure data collection includes the four core ABS standards at the minimum. Similarly, health administrative data collections used to inform research and service provision should align with these standards. One exception to this is research on racial discrimination where CALD standards may not be fit for use, and information indicating groups that have been ‘racialised’ may be more appropriate [68]. Second, using multiple indicators improves identification of CALD as single indicators provide a limited view of this heterogeneous group. We have demonstrated that using only country of birth resulted in a small proportion of children who were CALD in the census as many are born in Australia. Similarly, using ‘language spoken’ only may misclassify predominantly English speaking CALD children as non-CALD.

Identifying subgroups with greater need is crucial to progressive proportionate universal approaches for service delivery at population level [4]. It is important to examine the magnitude of health disparities within various subgroups of CALD. The patterning of health conditions may be similar to non-CALD groups for certain subgroups of CALD and not for others. Study analysis should be extended to assess the patterns of health outcomes of interest according to meaningful subcategories of CALD contingent on the research question. For example, a study on health literacy may assess CALD subgroups according to English language proficiency and year of arrival into Australia which may indicate ability to develop local health literacy. Conversely, a study examining the mental health impact of racial discrimination would assess subgroups according to parental country of birth as proxy for the concept of visible minority as demonstrated by Priest et al [80]. The points outlined above aspire towards a more consistent approach to best ascertain CALD children in Australian health research.

## Conclusion

Notwithstanding complexities in operationalising the concept of CALD, it is understood that inequities in health and wellbeing exists in people not belonging to the mainstream non-Indigenous population in Australia. Overall, there are not many published studies on CALD children’s health in Australia and the existing literature has a heavy focus on children of refugee background. Although studies that use the term CALD adhere to the ABS standards on cultural and language diversity to varying degrees, the overall approach to ascertaining CALD or migrant children is inconsistent. As such there can be large variation to the estimated size of the CALD child population which in turn will impact estimates of the health outcomes of interest and the inequities in those outcomes. If CALD children are indeed a priority population in the national action plan, more attention is required for a carefully considered and consistent approach to identifying these children in Australian health research. Although we cannot capture all the ways in which we are ethno-culturally different, we can certainly do better.

## Data Availability

The Australia Housing and Population Census 2016 and Australian Census and Migration Intergrated Dataset 2016 data are available, upon user registration and approval, on the website of the Australian Bureau of Statistics.

https://www.abs.gov.au/

## Supporting information

Table S1. CALD definition by various Australian government and non-government organisations

Table S2. Permanent visa categories for children and youth aged 0-19 years, ACMID 2016 (N = 386,820) & Census 2016 (N = 5,786,205)

Table S3. List of child health and wellbeing publications categorised according to how CALD was defined

Table S4. Australian child health research published since 2010 with focus on CALD, ethnic minority or migrants categorised into themes

Table S5. Australian child health research published since 2010 with focus on refugees and asylum seekers categorised into themes

