## Supporting material Tables S1-S5 for "Capturing cultural and linguistic diversity in child health research in Australia"

Table S1: CALD definition by various Australian government and non-government organisations

| **Organisation** | **Source** | **CALD definition** |
| --- | --- | --- |
| Federation of Ethnic Communities Council of Australia (FECCA) | Federation of Ethnic Communities Council of Australia (2015). Review of Australian Research on Older People from Culturally and Linguistically Diverse Backgrounds. Curtin ACT. | Born in a non-English speaking country |
| Australian Institute of Health and Welfare (AIHW) | Thow, A.M. & Waters, A-M. (2005). Diabetes in culturally and linguistically diverse Australians. Canberra: Australian Institute of Health and Welfare. | i) Main language spoken being other than English ii) Being born in countries where English is not the main language |
| Commonwealth Department of Social Services | Department of Social Services. (2015). National Ageing and Aged Care Strategy For people from Culturally and Linguistically Diverse (CALD) backgrounds. Commonwealth of Australia. | People from non-English speaking backgrounds |
|  | Department of Social Services. (2017). Multicultural Access and Equity in Australian Government Services Report 2013–2015. In Department of Social Services (Ed.): Commonwealth of Australia. | Australia’s non-Aboriginal and Torres Strait Islander cultural groups who are not from an English-speaking, Anglo-Celtic background |
| Office of Multicultural Interests (Western Australia) | Office of Multicultural Interests. (2010). Implementing the Principles of Multiculturalism Locally. West Perth: Department of Local Government. | Australia’s non-Aboriginal and Torres Strait Islander cultural groups who are not from an English-speaking, Anglo-Celtic background |

Table S2. Permanent visa categories for children and youth aged 0-19 years, ACMID 2016 (N = 386,820) & Census 2016 (N = 5,786,205)

| **Permanent Visa Type** | **Individuals aged 0 – 19 years in ACMID 2016**  **(N)** | **Proportion of migrant children and youth aged 0-19 years (ACMID 2016)** | **As a proportion of all children and youth aged 0-19 years in Australia (Census 2016)** |
| --- | --- | --- | --- |
| Skilled migration | 262,966 | 68% | 5% |
| Family | 68,566 | 18% | 1% |
| Humanitarian | 55,169 | 14% | 1% |
| Other Permanent | 119 | 0% | 0% |
| Total | 386,820 | 100% | 7% |

Table S3: List of child health and wellbeing publications categorised according to how CALD was defined

**CALD defined by language spoken other than English**

Barnett, L.M., Telford, R.M., Strugnell, C., Rudd, J., Olive, L.S., & Telford, R.D. (2019). Impact of cultural background on fundamental movement skill and its correlates. Journal of Sports Sciences, 37, 492-499.

Brown, J., Burton, D., Nikolin, S., Crooks, P.J., Hatfield, J., & Bilston, L.E. (2013). A qualitative approach using the integrative model of behaviour change to identify intervention strategies to increase optimal child restraint practices among culturally and linguistically diverse families in New South Wales. Injury Prevention, 19, 6-12.

Dudley, D.A., Okely, A.D., Pearson, P., Cotton, W.G., & Caputi, P. (2012). Changes in physical activity levels, lesson context, and teacher interaction during physical education in culturally and linguistically diverse Australian schools. Int J Behav Nutr Phys Act, 9, 114.

Guajardo, M.G.U., Kelly, C., Bond, K., Thomson, R., & Slewa-Younan, S. (2019). An evaluation of the teen and Youth Mental Health First Aid training with a CALD focus: an uncontrolled pilot study with adolescents and adults in Australia. International Journal of Mental Health Systems, 13.

Guo, X.Y., Woolfenden, S., McDonald, G., Saavedra, A., & Lingam, R. (2019). Discharge against medical advice in culturally and linguistically diverse Australian children. Archives of Disease in Childhood, 104, 1150-1154.

Hardy, L.L., Jin, K., Mihrshahi, S., & Ding, D. (2019). Trends in overweight, obesity, and waist-to-height ratio among Australian children from linguistically diverse backgrounds, 1997 to 2015. International Journal of Obesity, 43, 116-124.

Hardy, L.L., King, L., Hector, D., & Baur, L.A. (2013). Socio-cultural differences in Australian primary school children’s weight and weight-related behaviours. Journal of Paediatrics and Child Health, 49, 8.

Scott, B., Bolton, K.A., Strugnell, C., Allender, S., & Marks, J. (2019). Weight status and obesity-related dietary behaviours among culturally and linguistically diverse (CALD) children in Victoria, Australia. BMC Pediatr*,* 19, 511.

Sterman, J.J., Naughton, G.A., Bundy, A.C., Froude, E., & Villeneuve, M.A. (2019). Mothers supporting play as a choice for children with disabilities within a culturally and linguistically diverse community. Scand J Occup Ther, 1-12.

Wen, L.M., Orr, N., & Rissel, C. (2007). The role of ethnicity in determining access to and acceptability of home visiting for early childhood health and wellbeing. Aust Health Rev, 31, 132-139.

Woolfenden, S., Posada, N., Krchnakova, R., Crawford, J., Gilbert, J., Jursik, B., et al. (2015). Equitable access to developmental surveillance and early intervention - understanding the barriers for children from culturally and linguistically diverse (CALD) backgrounds. Health Expectations, 18, 3286-3301.

**CALD defined by language spoken combined with other variables such as country of birth, English proficiency, length of stay in Australia, religion and visa type**

Cyril, S., Green, J., Nicholson, J.M., Agho, K., & Renzaho, A.M. (2016). Exploring Service Providers' Perspectives in Improving Childhood Obesity Prevention among CALD Communities in Victoria, Australia. PLoS ONE, 11, e0162184.

Cyril, S., Nicholson, J.M., Agho, K., Polonsky, M., & Renzaho, A.M. (2017). Barriers and facilitators to childhood obesity prevention among culturally and linguistically diverse (CALD) communities in Victoria, Australia. Australian and New Zealand Journal of Public Health, 41, 287-293.

Garg, P., Ha, M.T., Eastwood, J., Harvey, S., Woolfenden, S., Murphy, E., et al. (2017). Explaining culturally and linguistically diverse (CALD) parents’ access of healthcare services for developmental surveillance and anticipatory guidance: qualitative findings from the ‘Watch Me Grow’ study. BMC Health Services Research, 17.

Khawaja, N.G., Allan, E., & Schweitzer, R.D. (2018). The Role of School Connectedness and Social Support in the Acculturation in Culturally and Linguistically Diverse Youth in Australia. Australian Psychologist, 53, 355-364.

Khawaja, N.G., & Carr, K. (2019). Exploring the factor structure and psychometric properties of an acculturation and resilience scale with culturally and linguistically diverse adolescents. Australian Psychologist.

Khawaja, N.G., Ibrahim, O., & Schweitzer, R.D. (2017). Mental Wellbeing of Students from Refugee and Migrant Backgrounds: The Mediating Role of Resilience. School Mental Health, 9, 284-293.

Khawaja, N.G., Pekin, C., & Schweitzer, R.D. (2019). Factor structure and psychometric properties of the Hopkins Symptom Checklist: An investigation with culturally and linguistically diverse youth in Australia. Australian Journal of Psychology, 71, 137-145.

Khawaja, N.G., & Ramirez, E. (2019). Building Resilience in Transcultural Adolescents: an Evaluation of a Group Program. Journal of Child and Family Studies, 28, 2977-2987.

Mitchelson, M.R., Erskine, H.E., Ramirez, E., Suleman, F., Prasad-Ildes, R., Siskind, D., et al. (2010). BRiTA Futures: A resilience-building program for children and young people from culturally and linguistically diverse backgrounds - Program description and preliminary findings. Advances in Mental Health*,* 9, 243-254.

Nicholson, J.M., Cann, W., Matthews, J., Berthelsen, D., Ukoumunne, O.C., Trajanovska, M., et al. (2016). Enhancing the early home learning environment through a brief group parenting intervention: study protocol for a cluster randomised controlled trial. BMC Pediatrics, 16.

O'Connor, M., Slopen, N., Becares, L., Burgner, D., Williams, D.R., & Priest, N. (2019). Inequalities in the Distribution of Childhood Adversity From Birth to 11 Years. Acad Pediatr.

Priest, N., Baxter, J., & Hayes, L. (2012). Social and emotional outcomes of Australian children from Indigenous and culturally and linguistically diverse backgrounds. Australian and New Zealand Journal of Public Health, 36, 183-190.

Rosso, E., & McGrath, R. (2016). Promoting physical activity among children and youth in disadvantaged South Australian CALD communities through alternative community sport opportunities. Health Promot J Austr, 27, 105-110.

Scrafton, E., & Whitington, V. (2015). The accessibility of socio-dramatic play to culturally and linguistically diverse Australian preschoolers. European Early Childhood Education Research Journal, 23, 213-228.

White, L., & Chalmers, S. (2011). Responding to cultural diversity at two Sydney-based children's hospitals. Journal of Paediatrics and Child Health, 47, 788-794.

**CALD defined by being born overseas or born in a non-English speaking country**

Ogbo, F.A., Eastwood, J., Page, A., Arora, A., McKenzie, A., Jalaludin, B., et al. (2016). Prevalence and determinants of cessation of exclusive breastfeeding in the early postnatal period in Sydney, Australia. Int Breastfeed J, 12, 16.

Sawrikar, P. (2013). A qualitative study on the pros and cons of ethnically matching culturally and linguistically diverse (CALD) client families and child protection caseworkers. Children and Youth Services Review, 35, 321-331.

Strugnell, C., A, M.N.R., Ridley, K., & Burns, C. (2015). Physical activity and sedentary behaviour among Asian and Anglo-Australian adolescents. Health Promot J Austr, 26, 105-114.

Strugnell, C., Renzaho, A., Ridley, K., & Burns, C. (2011). Reliability of the modified child and adolescent physical activity and nutrition survey, physical activity (CAPANS-PA) questionnaire among Chinese-Australian youth. BMC Med Res Methodol, 11, 122.

Taft, A.J., Small, R., Hegarty, K.L., Watson, L.F., Gold, L., & Lumley, J.A. (2011). Mothers' AdvocateS In the Community (MOSAIC)-non-professional mentor support to reduce intimate partner violence and depression in mothers: a cluster randomised trial in primary care. Bmc Public Health, 11.

von Katterfeld, B., Li, J., McNamara, B., & Langridge, A.T. (2012). Maternal and neonatal outcomes associated with gestational diabetes in women from culturally and linguistically diverse backgrounds in Western Australia. Diabet Med, 29, 372-377.

**CALD defined by ‘cultural group’ or ‘background’**

Deans, J., Liang, R., & Frydenberg, E. (2016). Giving voices and providing skills to families in culturally and linguistically diverse communities through a productive parenting program. Australasian Journal of Early Childhood, 41, 13-18.

Frydenberg, E., Deans, J., & Liang, R. (2019). Parents and Children's Coping: Building Resilience and Wellbeing in the Early Years. Social Indicators Research, 145, 629-640.

Sepulveda, M., Henderson, S., Farrell, D., & Heuft, G. (2016). Needs-gap analysis on culturally and linguistically diverse grandparent carers' 'hidden issues': a quality improvement project. Australian Journal of Primary Health, 22, 477-482.

**CALD defined by directly questioning study participants if they identify as CALD**

Ogbo, F.A., Ezeh, O.K., Dhami, M.V., Naz, S., Khanlari, S., McKenzie, A., et al. (2019a). Perinatal Distress and Depression in Culturally and Linguistically Diverse (CALD) Australian Women: The Role of Psychosocial and Obstetric Factors. International Journal of Environmental Research and Public Health, 16.

Ogbo, F.A., Ezeh, O.K., Khanlari, S., Naz, S., Senanayake, P., Ahmed, K.Y., et al. (2019b). Determinants of Exclusive Breastfeeding Cessation in the Early Postnatal Period among Culturally and Linguistically Diverse (CALD) Australian Mothers. Nutrients, 11.

Table S4: Australian child health research published since 2010 with focus on CALD, ethnic minority or migrants categorised into themes

**List of publications**

*Theme: Chronic disease & lifestyle risk factors*

Scott, B., et al. (2019). "Weight status and obesity-related dietary behaviours among culturally and linguistically diverse (CALD) children in Victoria, Australia." BMC Pediatr **19**(1): 511.

Barnett, L. M., et al. (2019). "Impact of cultural background on fundamental movement skill and its correlates." Journal of Sports Sciences **37**(5): 492-499.

Hardy, L. L., et al. (2019). "Trends in overweight, obesity, and waist-to-height ratio among Australian children from linguistically diverse backgrounds, 1997 to 2015." International Journal of Obesity **43**(1): 116-124.

Cyril, S., et al. (2017). "Barriers and facilitators to childhood obesity prevention among culturally and linguistically diverse (CALD) communities in Victoria, Australia." Aust N Z J Public Health **41**(3): 287-293.

Okely, A. D., et al. (2017). "Promoting motor skills in low-income, ethnic children: The Physical Activity in Linguistically Diverse Communities (PALDC) nonrandomized trial." J Sci Med Sport **20**(11): 1008-1014.

Cyril, S., et al. (2016). "Exploring Service Providers' Perspectives in Improving Childhood Obesity Prevention among CALD Communities in Victoria, Australia." PLoS ONE **11**(10): e0162184.

Cyril, S., et al. (2016). "Relationship between body mass index and family functioning, family communication, family type and parenting style among African migrant parents and children in Victoria, Australia: a parent-child dyad study." BMC Public Health **15**: 707.

Renzaho, A. M., et al. (2015). "The Healthy Migrant Families Initiative: development of a culturally competent obesity prevention intervention for African migrants." BMC Public Health **15**: 272.

Strugnell, C., et al. (2015). "Physical activity and sedentary behaviour among Asian and Anglo-Australian adolescents." Health Promot J Austr **26**(2): 105-114.

Strugnell, C., et al. (2014). "Reliability and validity of the modified Child and Adolescent Physical Activity and Nutrition Survey (CAPANS-C) questionnaire examining potential correlates of physical activity participation among Chinese-Australian youth." BMC Public Health **14**: 145.

Griffith, M., et al. (2014). "Migration-related influences on obesity among sub-Saharan African migrant adolescents in Melbourne, Australia." Nutrition & Dietetics **71**(4): 252-257.

Hardy, L. L., et al. (2013). "Socio-cultural differences in Australian primary school children’s weight and weight-related behaviours." Journal of Paediatrics and Child Health **49**: 8.

Dudley, D. A., et al. (2012). "Changes in physical activity levels, lesson context, and teacher interaction during physical education in culturally and linguistically diverse Australian schools." Int J Behav Nutr Phys Act **9**: 114.

Strugnell, C., et al. (2011). "Reliability of the modified child and adolescent physical activity and nutrition survey, physical activity (CAPANS-PA) questionnaire among Chinese-Australian youth." BMC Med Res Methodol **11**: 122.

*Theme: Access and utilization of health services*

Guo, X. Y., et al. (2019). "Discharge against medical advice in culturally and linguistically diverse Australian children." Arch Dis Child **104**(12): 1150-1154.

Garg, P., et al. (2017). "Explaining culturally and linguistically diverse (CALD) parents' access of healthcare services for developmental surveillance and anticipatory guidance: qualitative findings from the 'Watch Me Grow' study." BMC Health Serv Res **17**(1): 228.

Botfield, J. R., et al. (2017). "Drawing them in: professional perspectives on the complexities of engaging "culturally diverse' young people with sexual and reproductive health promotion and care in Sydney, Australia." Culture Health & Sexuality **19**(4): 438-452.

Woolfenden, S., et al. (2015). "Equitable access to developmental surveillance and early intervention--understanding the barriers for children from culturally and linguistically diverse (CALD) backgrounds." Health Expect **18**(6): 3286-3301.

Christian, B., et al. (2015). "Exploring child dental service use among migrant families in metropolitan Melbourne, Australia." Australian Dental Journal **60**(2): 200-204.

Riggs, E., et al. (2014). "Hard to reach communities or hard to access services? Migrant mothers' experiences of dental services." Australian Dental Journal **59**(2): 201-207.

Sims, M., et al. (2014). "Inclusive Services for Children and Families From CaLD Backgrounds in an Australian Context." Sage Open **4**(1).

Lai, F. Y., et al. (2014). "Examining potential barriers to early intervention access in Australian hearing impaired children." Int J Pediatr Otorhinolaryngol **78**(3): 507-512.

White, L. and S. Chalmers (2011). "Responding to cultural diversity at two Sydney-based children's hospitals." J Paediatr Child Health **47**(11): 788-794.

*Theme: Maternal & neonatal health outcomes*

Ogbo, F. A., et al. (2019). "Perinatal Distress and Depression in Culturally and Linguistically Diverse (CALD) Australian Women: The Role of Psychosocial and Obstetric Factors." Int J Environ Res Public Health **16**(16).

Ogbo, F. A., et al. (2019). "Determinants of Exclusive Breastfeeding Cessation in the Early Postnatal Period among Culturally and Linguistically Diverse (CALD) Australian Mothers." Nutrients **11**(7).

Ogbo, F. A., et al. (2016). "Prevalence and determinants of cessation of exclusive breastfeeding in the early postnatal period in Sydney, Australia." Int Breastfeed J **12**: 16.

von Katterfeld, B., et al. (2012). "Maternal and neonatal outcomes associated with gestational diabetes in women from culturally and linguistically diverse backgrounds in Western Australia." Diabet Med **29**(3): 372-377.

Taft, A. J., et al. (2011). "Mothers' AdvocateS In the Community (MOSAIC)-non-professional mentor support to reduce intimate partner violence and depression in mothers: a cluster randomised trial in primary care." BMC Public Health **11**.

*Theme: Parenting practices*

Sterman, J. J., et al. (2019). "Mothers supporting play as a choice for children with disabilities within a culturally and linguistically diverse community." Scand J Occup Ther: 1-12.

Frydenberg, E., et al. (2019). "Parents and Children's Coping: Building Resilience and Wellbeing in the Early Years." Social Indicators Research **145**(2): 629-640.

Hall, A., et al. (2018). "Barriers to correct child restraint use: A qualitative study of child restraint users and their needs." Safety Science **109**: 186-194.

Deans, J., et al. (2016). "Giving voices and providing skills to families in culturally and linguistically diverse communities through a productive parenting program." Australasian Journal of Early Childhood **41**(1): 13-18.

Sepulveda, M., et al. (2016). "Needs-gap analysis on culturally and linguistically diverse grandparent carers' 'hidden issues': a quality improvement project." Aust J Prim Health **22**(6): 477-482.

Nicholson, J. M., et al. (2016). "Enhancing the early home learning environment through a brief group parenting intervention: study protocol for a cluster randomised controlled trial." BMC Pediatrics **16**.

Brown, J., et al. (2013). "A qualitative approach using the integrative model of behaviour change to identify intervention strategies to increase optimal child restraint practices among culturally and linguistically diverse families in New South Wales." Injury Prevention **19**(1): 6-12.

*Theme: Mental health & wellbeing*

Uribe Guajardo, M. G., et al. (2019). "An evaluation of the teen and Youth Mental Health First Aid training with a CALD focus: an uncontrolled pilot study with adolescents and adults in Australia." Int J Ment Health Syst **13**: 73.

Khawaja, N. G. and Ramirez, E. (2019). "Building Resilience in Transcultural Adolescents: an Evaluation of a Group Program." Journal of Child and Family Studies **28**(11): 2977-2987.

Khawaja, N. G. and Carr, K. (2019). "Exploring the factor structure and psychometric properties of an acculturation and resilience scale with culturally and linguistically diverse adolescents." Australian Psychologist **55**(1): 26-37.

Khawaja, N. G., et al. (2019). "Factor structure and psychometric properties of the Hopkins Symptom Checklist: An investigation with culturally and linguistically diverse youth in Australia." Australian Journal of Psychology **71**(2): 137-145.

Khawaja, N. G., et al. (2018). "The Role of School Connectedness and Social Support in the Acculturation in Culturally and Linguistically Diverse Youth in Australia." Australian Psychologist **53**(4): 355-364.

Khawaja, N. G., et al. (2017). "Mental Wellbeing of Students from Refugee and Migrant Backgrounds: The Mediating Role of Resilience." School Mental Health **9**(3): 284-293.

Scrafton, E. and V. Whitington (2015). "The accessibility of socio-dramatic play to culturally and linguistically diverse Australian preschoolers." European Early Childhood Education Research Journal **23**(2): 213-228.

Priest, N., et al. (2012). "Social and emotional outcomes of Australian children from Indigenous and culturally and linguistically diverse backgrounds." Aust N Z J Public Health **36**(2): 183-190.

Mitchelson, M. R., et al. (2010). "BRiTA Futures: A resilience-building program for children and young people from culturally and linguistically diverse backgrounds - Program description and preliminary findings." Advances in Mental Health **9**(3): 243-254.

*Theme: Child maltreatment & childhood adversity*

O'Connor M, et al. (2019) Inequalities in the Distribution of Childhood Adversity From Birth to 11 Years. Acad Pediatr S1876-2859(19)30506-6.

Waniganayake, M., et al. (2019). "Maintaining culture and supporting cultural identity in foster care placements." Australasian Journal of Early Childhood **44**(4): 365-377.

Sawrikar, P. (2013). "A qualitative study on the pros and cons of ethnically matching culturally and linguistically diverse (CALD) client families and child protection caseworkers." Children and Youth Services Review **35**(2): 321-331.

Sawrikar, P. (2014). "Inadequate supervision or inadequate sensitivity to cultural differences in parenting? Exploring cross-cultural rates of neglect in an Australian sample." Qualitative Social Work **13**(5): 619-635.

Sawrikar, P. and I. Katz (2014). "‘Normalizing the Novel’: How Is Culture Addressed in Child Protection Work With Ethnic-Minority Families in Australia?" Journal of Social Service Research **40**(1): 39-61.

Zurynski, Y., et al. (2017). "Female genital mutilation in children presenting to Australian paediatricians." Archives of Disease in Childhood **102**(6): 509-515.

*Theme: Oral health disease*

Arora, A., et al. (2018). "Adaptation of child oral health education leaflets for Arabic migrants in Australia: a qualitative study." BMC Oral Health **18**(1): 10.

Gibbs, L., et al. (2016). "Child oral health in migrant families: A cross-sectional study of caries in 1-4 year old children from migrant backgrounds residing in Melbourne, Australia." Community Dent Health **33**(2): 100-106.

Gibbs, L., et al. (2015). "Teeth Tales: a community-based child oral health promotion trial with migrant families in Australia." BMJ Open **5**(6).

Riggs, E., et al. (2015). "Breaking down the barriers: a qualitative study to understand child oral health in refugee and migrant communities in Australia." Ethnicity & Health **20**(3): 241-257.

Riggs, E., et al. (2014). "Assessing the cultural competence of oral health research conducted with migrant children." Community Dent Oral Epidemiol **42**(1): 43-52.

Arora, A., et al. (2014). "'What do these words mean?': A qualitative approach to explore oral health literacy in Vietnamese immigrant mothers in Australia." Health Education Journal **73**(3): 303-312.

*Theme: Discrimination & health effects*

Priest, N., et al. (2019). "Cumulative Effects of Bullying and Racial Discrimination on Adolescent Health in Australia." Journal of Health and Social Behavior **60**(3): 344-361.

Priest, N., et al. (2016). "Bullying Victimization and Racial Discrimination Among Australian Children." Am J Public Health **106**(10): 1882-1884.

Priest, N., et al. (2014). "Experiences of racism, racial/ethnic attitudes, motivated fairness and mental health outcomes among primary and secondary school students." J Youth Adolesc **43**(10): 1672-1687.

*Theme: Physical health*

Paxton, G. A., et al. (2016). "No Jab, No Pay - no planning for migrant children." Med J Aust **205**(7): 296-298.

Munns, C. F., et al. (2012). "Incidence of vitamin D deficiency rickets among Australian children: an Australian Paediatric Surveillance Unit study." Med J Aust **196**(7): 466-468.

Paxton, G. A., et al. (2011). "East African immigrant children in Australia have poor immunisation coverage." J Paediatr Child Health **47**(12): 888-892.

*Theme: Disabilities*

Abdullahi, I., et al. (2019). "Hospital admissions in children with developmental disabilities from ethnic minority backgrounds." Dev Med Child Neurol.

Fulcher, A. N., et al. (2015). "Factors influencing speech and language outcomes of children with early identified severe/profound hearing loss: Clinician-identified facilitators and barriers." Int J Speech Lang Pathol **17**(3): 325-333.

Ziviani, J., et al. (2014). "Early intervention services of children with physical disabilities: complexity of child and family needs." Aust Occup Ther J **61**(2): 67-75.

Table S5: Australian child health research published since 2010 with focus on refugees and asylum seekers categorised into themes

**List of publications**

*Theme: General health profile*

Hanes, G., et al. (2019). "Paediatric asylum seekers in Western Australia: Identification of adversity and complex needs through comprehensive refugee health assessment." J Paediatr Child Health **55**(11): 1367-1373.

Hirani, K., et al. (2019). "Medical needs of adolescent refugees resettling in Western Australia." Arch Dis Child **104**(9): 880-883.

Heenan, R. C., et al. (2019). "'I think we've had a health screen': New offshore screening, new refugee health guidelines, new Syrian and Iraqi cohorts: Recommendations, reality, results and review." J Paediatr Child Health **55**(1): 95-103.

Zwi, K., et al. (2018). "Helping refugee children thrive: what we know and where to next." Arch Dis Child **103**(6): 529-532.

Zwi, K., et al. (2017). "Refugee children and their health, development and well-being over the first year of settlement: A longitudinal study." J Paediatr Child Health **53**(9): 841-849.

Zwi, K., et al. (2016). "Methods for a longitudinal cohort of refugee children in a regional community in Australia." BMJ Open **6**(8): e011387.

Hirani, K., et al. (2016). "Health of adolescent refugees resettling in high-income countries." Arch Dis Child **101**(7): 670-676.

Mutch, R. C., et al. (2012). "Tertiary paediatric refugee health clinic in Western Australia: analysis of the first 1026 children." J Paediatr Child Health **48**(7): 582-587.

Giallo, R., et al. (2017). "The physical and mental health problems of refugee and migrant fathers: findings from an Australian population-based study of children and their families." BMJ Open **7**(11): e015603.

Refugee child health checks. (2011) Aust Nurs J;18(10): 43.

*Theme: Mental health and wellbeing*

Baak, M., et al. (2020). "The Role of Schools in Identifying and Referring Refugee Background Young People Who Are Experiencing Mental Health Issues." J Sch Health **90**(3):172-181.

Lamb, C. S. (2020). "Constructing early childhood services as culturally credible trauma-recovery environments: participatory barriers and enablers for refugee families." European Early Childhood Education Research Journal **28**(1): 129-148.

Baker, J. R., et al. (2019). "Optimising refugee children's health/wellbeing in preparation for primary and secondary school: a qualitative inquiry." Bmc Public Health **19**(1): 812.

de Anstiss, H., et al. (2019). "Relationships in a new country: A qualitative study of the social connections of refugee youth resettled in South Australia." Journal of Youth Studies **22**(3): 346-362.

Due, C., et al. (2019). ""At night he cries from dreams": Perceptions of children's psychological distress and wellbeing amongst parents with refugee or asylum seeker backgrounds in Australia." Australian Psychologist **54**(5): 438-449.

Essex, R. (2019). "The psychometric properties of the strengths and difficulties questionnaire for children from refugee backgrounds in Australia." Clinical Psychologist **23**(3): 261-270.

Lawrence, J. A., et al. (2019). "Perspectives of Refugee Children Resettling in Australia on Indicators of Their Wellbeing." Child Indicators Research **12**(3): 943-962.

Bennouna, C., et al. (2019). "School-based programs for Supporting the mental health and psychosocial wellbeing of adolescent forced migrants in high-income countries: A scoping review." Social Science & Medicine **239**.

Reid, K. and D. Berle (2019). "Parental trajectories of PTSD and child adjustment: Findings from the Building a New Life in Australia study." Am J Orthopsychiatry.

Mom, S., et al. (2019). "Capoeira Angola: An alternative intervention program for traumatized adolescent refugees from war-torn countries." Torture **29**(1): 85-96.

Cleary, M., et al. (2019). "Mental Health of Refugee Children: A Discursive Look at Causes, Considerations and Interventions." Issues Ment Health Nurs **40**(8): 665-671.

Bryant, R. A., et al. (2018). "The effect of post-traumatic stress disorder on refugees' parenting and their children's mental health: a cohort study." Lancet Public Health **3**(5): e249-e258.

Cameron, G., et al. (2018). "How Young Refugees Cope with Conflict in Culturally and Linguistically Diverse Urban Schools." Australian Psychologist **53**(2): 171-180.

Hirani, K., et al. (2018). "Identification of health risk behaviours among adolescent refugees resettling in Western Australia." Arch Dis Child **103**(3): 240-246.

Lau, W., et al. (2018). "Adjustment of refugee children and adolescents in Australia: outcomes from wave three of the Building a New Life in Australia study." BMC Med **16**(1): 157.

Momartin, S., et al. (2018). "Resilience building through alternative intervention: 'STARTTS "Project Bantu Capoeira Angola"'; on the road to recovery." Intervention-International Journal of Mental Health Psychosocial Work and Counselling in Areas of Armed Conflict **16**(2): 154-160.

Ratnamohan, L., et al. (2018). "Ghosts, tigers and landmines in the nursery: Attachment narratives of loss in Tamil refugee children with dead or missing fathers." Clin Child Psychol Psychiatry **23**(2): 294-310.

Tozer, M., et al. (2018). "Protective Factors Contributing to Wellbeing Among Refugee Youth in Australia." Journal of Psychologists and Counsellors in Schools **28**(1): 66-83.

Ziaian, T., et al. (2018). "Refugee Students' Psychological Wellbeing and Experiences in the Australian Education System: A Mixed-methods Investigation." Australian Psychologist **53**(4): 345-354.

Zwi, K., et al. (2018). "Protective factors for social-emotional well-being of refugee children in the first three years of settlement in Australia." Arch Dis Child **103**(3): 261-268.

Zwi, K., et al. (2018). "The impact of detention on the social-emotional wellbeing of children seeking asylum: a comparison with community-based children." Eur Child Adolesc Psychiatry **27**(4): 411-422.

Buchanan, Z. E., et al. (2017). "The Interconnection between Acculturation and Subjective and Social Wellbeing among Refugee Youth in Australia." Journal of Refugee Studies **30**(4): 511-529.

Essex, R. and P. Govintharajah (2017). "Mental health of children and adolescents in Australian alternate places of immigration detention." J Paediatr Child Health **53**(6): 525-528.

Hanes, G., et al. (2017). "Adversity and resilience amongst resettling Western Australian paediatric refugees." J Paediatr Child Health **53**(9): 882-888.

Riggs, D. W., et al. (2017). "'I Want to Bring Him from the Aeroplane to Here': The Meaning of Animals to Children of Refugee or Migrant Backgrounds Resettled in Australia." Children & Society **31**(3): 219-230.

Mares, S. (2016). "The Mental Health of Children and Parents Detained on Christmas Island: Secondary Analysis of an Australian Human Rights Commission Data Set." Health Hum Rights **18**(2): 219-232.

McGregor, L. S., et al. (2016). "An exploration of the adaptation and development after persecution and trauma (ADAPT) model with resettled refugee adolescents in Australia: A qualitative study." Transcultural Psychiatry **53**(3): 347-367.

Ooi, C. S., et al. (2016). "The Efficacy of a Group Cognitive Behavioral Therapy for War-Affected Young Migrants Living in Australia: A Cluster Randomized Controlled Trial." Frontiers in Psychology **7**: 1641.

Correa-Velez, I., et al. (2015). "The persistence of predictors of wellbeing among refugee youth eight years after resettlement in Melbourne, Australia." Social Science & Medicine **142**: 163-168.

McGregor, L. S., et al. (2015). "Familial separations, coping styles, and PTSD symptomatology in resettled refugee youth." J Nerv Ment Dis **203**(6): 431-438.

Mares, S. and K. Zwi (2015). "Sadness and fear: The experiences of children and families in remote Australian immigration detention." J Paediatr Child Health **51**(7): 663-669.

Posselt, M., et al. (2015). "Merging perspectives: obstacles to recovery for youth from refugee backgrounds with comorbidity." Australas Psychiatry **23**(3): 293-299.

Posselt, M., et al. (2015). "Aetiology of Coexisting Mental Health and Alcohol and Other Drug Disorders: Perspectives of Refugee Youth and Service Providers." Australian Psychologist **50**(2): 130-140.

Mace, A. O., et al. (2014). "Educational, developmental and psychological outcomes of resettled refugee children in Western Australia: a review of School of Special Educational Needs: Medical and Mental Health input." J Paediatr Child Health **50**(12): 985-992.

Ziaian, T., et al. (2013). "Emotional and Behavioural Problems Among Refugee Children and Adolescents Living in South Australia." Australian Psychologist **48**(2): 139-148.

Ziaian, T., et al. (2012). "Depressive symptomatology and service utilisation among refugee children and adolescents living in South Australia." Child and Adolescent Mental Health **17**(3): 146-152.

Henley, J. and J. Robinson (2011). "Mental health issues among refugee children and adolescents." Clinical Psychologist **15**(2): 51-62.

*Theme: Physical health (specific topics)*

Ghosh, S., et al. (2020). "Tuberculosis infection in under-2-year-old refugees: Should we be screening? A systematic review and meta-regression analysis." J Paediatr Child Health **56**(4): 622-629.

Colgan, K., et al. (2019). "Latent tuberculosis may be missed by current screening practices: Analysis of interferon-gamma release assay results from a paediatric refugee clinic." J Paediatr Child Health **55**(7): 826-832.

Volkman, T., et al. (2019). "Interpretation and management of discordant tuberculin skin test and interferon-gamma release assays results in children." J Paediatr Child Health **55**(2): 247-248.

Newman, K., et al. (2019). "Nutritional assessment of resettled paediatric refugees in Western Australia." J Paediatr Child Health **55**(5): 574-581.

Elliot, C., et al. (2018). "Tuberculin skin test versus interferon-gamma release assay in refugee children: A retrospective cohort study." J Paediatr Child Health **54**(8): 834-839.

Wadia, U., et al. (2018). "Randomised Controlled Trial Comparing Daily Versus Depot Vitamin D3 Therapy in 0-16-Year-Old Newly Settled Refugees in Western Australia Over a Period of 40 Weeks." Nutrients **10**(3).

Zurynski, Y., et al. (2017). "Female genital mutilation in children presenting to Australian paediatricians." Archives of Disease in Childhood **102**(6): 509-515.

Isaacs, D. and B. Tobin (2017). "Female genital mutilation and the role of health-care practitioners." J Paediatr Child Health **53**(6): 523-524.

Benson, J., et al. (2017). "Newly arrived refugee children with Helicobacter pylori are thinner than their non-infected counterparts." Aust J Prim Health **23**(1): 92-96.

Chuang, S. Y., et al. (2017). "An unusual case of haemolytic anaemia and failure to thrive in a Burmese refugee baby." J Paediatr Child Health **53**(5): 500-502.

Vaska, A. I., et al. (2016). "Age determination in refugee children: A narrative history tool for use in holistic age assessment." J Paediatr Child Health **52**(5): 523-528.

Sypek, S. A., et al. (2016). "A holistic approach to age estimation in refugee children." J Paediatr Child Health **52**(6): 614-620.

Graham, H. R., et al. (2016). "Learning Problems in Children of Refugee Background: A Systematic Review." Pediatrics **137**(6).

Rosso, E. and R. McGrath (2016). "Promoting physical activity among children and youth in disadvantaged South Australian CALD communities through alternative community sport opportunities." Health Promot J Austr **27**(2): 105-110.

Marais, B. J. (2014). "Tuberculosis in children." J Paediatr Child Health **50**(10): 759-767.

Goldwater, P. N. (2013). "Iatrogenic blood-borne viral infections in refugee children from war and transition zones." Emerg Infect Dis **19**(6): 892-898.

Joshua, P. R., et al. (2013). "Australian population cohort study of newly arrived refugee children: how effective is predeparture measles and rubella vaccination?" Pediatr Infect Dis J **32**(2): 104-109.

McMichael, C. (2013). "Unplanned but not unwanted? Teen pregnancy and parenthood among young people with refugee backgrounds." Journal of Youth Studies **16**(5): 663-678.

Gray, K., et al. (2012). "Vitamin d and tuberculosis status in refugee children." Pediatr Infect Dis J **31**(5): 521-523.

Sheikh, M., et al. (2011). "Vitamin D deficiency in refugee children from conflict zones." J Immigr Minor Health **13**(1): 87-93.

Lucas, M., et al. (2010). "A prospective large-scale study of methods for the detection of latent Mycobacterium tuberculosis infection in refugee children." Thorax **65**(5): 442-448.

Cherian, S., et al. (2010). "Associations between Helicobacter pylori infection, co-morbid infections, gastrointestinal symptoms, and circulating cytokines in African children." Helicobacter **15**(2): 88-97.

*Theme: Oral health disease*

Patel, J., et al. (2019). "Infant oral mutilation: A case report and learning points." J Paediatr Child Health.

Riggs, E., et al. (2017). "Refugee child oral health." Oral Diseases **23**(3): 292-299.

Nicol, P., et al. (2015). "Caries burden and efficacy of a referral pathway in a cohort of preschool refugee children." Aust Dent J **60**(1): 73-79.

Quach, A., et al. (2015). "Gaps in smiles and services: a cross-sectional study of dental caries in refugee-background children." BMC Oral Health **15**: 10.

Nicol, P., et al. (2014). "Informing a culturally appropriate approach to oral health and dental care for pre-school refugee children: a community participatory study." BMC Oral Health **14**: 69.

*Theme: Social wellbeing associated with resettlement*

Lawrence, J. A., et al. (2019). "Perspectives of Refugee Children Resettling in Australia on Indicators of Their Wellbeing." Child Indicators Research **12**(3): 943-962.

de Anstiss, H., et al. (2019). "Relationships in a new country: A qualitative study of the social connections of refugee youth resettled in South Australia." Journal of Youth Studies **22**(3): 346-362.

Wong, C. W. S., et al. (2018). "Individual, Pre-Migration, and Post-Settlement Factors in Predicting Academic Success of Adolescents from Refugee Backgrounds: a 12-Month Follow-Up." Journal of International Migration and Integration **19**(4): 1095-1117.

Buchanan, Z. E., et al. (2018). "Perceived discrimination, language proficiencies, and adaptation: Comparisons between refugee and non-refugee immigrant youth in Australia." International Journal of Intercultural Relations **63**: 105-112.

Correa-Velez, I., et al. (2017). "Predictors of Secondary School Completion Among Refugee Youth 8 to 9 Years After Resettlement in Melbourne, Australia." Journal of International Migration and Integration **18**(3): 791-805.

Block, K. and L. Gibbs (2017). "Promoting Social Inclusion through Sport for Refugee-Background Youth in Australia: Analysing Different Participation Models." Social Inclusion **5**(2): 91-100.

Nardone, M. and I. Correa-Velez (2016). "Unpredictability, Invisibility and Vulnerability: Unaccompanied Asylum-Seeking Minors' Journeys to Australia." Journal of Refugee Studies **29**(3): 295-314.

Due, C., et al. (2016). "Experiences of School Belonging for Young Children With Refugee Backgrounds." Educational and Developmental Psychologist **33**: 33-53.

Losoncz, I. (2016). "Building safety around children in families from refugee backgrounds: Ensuring children's safety requires working in partnership with families and communities." Child Abuse Negl **51**: 416-426.

Correa-Velez, I., et al. (2015). "The persistence of predictors of wellbeing among refugee youth eight years after resettlement in Melbourne, Australia." Social Science & Medicine **142**: 163-168.

MacMillan, K. K., et al. (2015). "Refugee children's play: Before and after migration to Australia." J Paediatr Child Health **51**(8): 771-777.

Earnest, J., et al. (2015). "Resettlement experiences and resilience in refugee youth in Perth, Western Australia." BMC Res Notes **8**: 236.

Block, K., et al. (2014). "Supporting schools to create an inclusive environment for refugee students." International Journal of Inclusive Education **18**(12): 1337-1355.

Levi, M. (2014). "Mothering in transition: The experiences of Sudanese refugee women raising teenagers in Australia." Transcultural Psychiatry **51**(4): 479-498.

Schweitzer, R. D., et al. (2014). "Narratives of healing: A case study of a young Liberian refugee settled in Australia." Arts in Psychotherapy 41(1): 98-106.

Gifford, S. M. and R. Wilding (2013). "Digital Escapes? ICTs, Settlement and Belonging among Karen Youth in Melbourne, Australia." Journal of Refugee Studies 26(4): 558-575.

McMichael, C., et al. (2011). "Negotiating family, navigating resettlement: family connectedness amongst resettled youth with refugee backgrounds living in Melbourne, Australia." Journal of Youth Studies **14**(2): 179-195.

Correa-Velez, I., et al. (2010). "Longing to belong: social inclusion and wellbeing among youth with refugee backgrounds in the first three years in Melbourne, Australia." Soc Sci Med **71**(8): 1399-1408.

Sampson, R. and S. M. Gifford (2010). "Place-making, settlement and well-being: The therapeutic landscapes of recently arrived youth with refugee backgrounds." Health & Place **16**(1): 116-131.

*Theme: Access to services and service delivery models*

Yelland, J., et al. (2018). "Improving the ascertainment of refugee-background people in health datasets and health services." Australian Health Review **42**(2): 130-133.

Willey, S. M., et al. (2018). "Maternal and child health nurses work with refugee families: Perspectives from regional Victoria, Australia." J Clin Nurs **27**(17-18): 3387-3396.

Paxton, G. A., et al. (2018). "Catching up with catch-up: a policy analysis of immunisation for refugees and asylum seekers in Victoria." Aust J Prim Health **24**(6): 480-490.

Kiang, K. M., et al. (2018). "No jab, no record: Catch-up vaccination of children in immigration detention." J Paediatr Child Health **54**(4): 348-350.

Zwi, K., et al. (2017). "Screening and Primary Care Access for Newly Arrived Paediatric Refugees in Regional Australia: A 5 year Cross-sectional Analysis (2007-12)." J Trop Pediatr **63**(2): 109-117.

Nelson, D., et al. (2017). "Critical social work with unaccompanied asylum-seeking young people: Restoring hope, agency and meaning for the client and worker." International Social Work **60**(3): 601-613.

Posselt, M., et al. (2017). "Improving the provision of services to young people from refugee backgrounds with comorbid mental health and substance use problems: addressing the barriers." BMC Public Health **17**(1): 280.

Boylen, S., et al. (2017). "Impact of professional interpreters on outcomes of hospitalized children from migrant and refugee families with low English proficiency: a systematic review protocol." JBI Database System Rev Implement Rep **15**(2): 202-211.

Woodland, L., et al. (2016). "Evaluation of a school screening programme for young people from refugee backgrounds." J Paediatr Child Health **52**(1): 72-79.

Zwi, K., et al. (2015). "Prioritizing vulnerable children: strategies to address inequity." Child Care Health Dev **41**(6): 827-835.

Yelland, J., et al. (2015). "Bridging the Gap: using an interrupted time series design to evaluate systems reform addressing refugee maternal and child health inequalities." Implement Sci **10**: 62.

Yelland, J., et al. (2014). "How do Australian maternity and early childhood health services identify and respond to the settlement experience and social context of refugee background families?" BMC Pregnancy Childbirth **14**: 348.

Garakasha, N. (2014). "Working with refugee young people: a nurse's perspective." Australian Journal of Advanced Nursing **32**(2): 24-31.

Patradoon-Ho, P. S. and R. W. Ambler (2012). "Universal post-arrival screening for child refugees in Australia: isn't it time?" J Paediatr Child Health **48**(2): 99-102.

Riggs, E., et al. (2012). "Accessing maternal and child health services in Melbourne, Australia: Reflections from refugee families and service providers." BMC Health Services Research **12**.

Copley, J., et al. (2011). "Development and evaluation of an occupational therapy program for refugee high school students." Aust Occup Ther J **58**(4): 310-316.

Woodland, L., et al. (2010). "Health service delivery for newly arrived refugee children: a framework for good practice." J Paediatr Child Health **46**(10): 560-567.

De Anstiss, H. and T. Ziaian (2010). "Mental health help-seeking and refugee adolescents: Qualitative findings from a mixed-methods investigation." Australian Psychologist **45**(1): 29-37.

*Theme: Conduct of research on refugees*

Hirani, K., et al. (2019). "Complexities of conducting research in adolescent refugees resettling in Australia." Journal of Paediatrics and Child Health **55**(8): 890-894.

Zwi, K., et al. (2016). "Methods for a longitudinal cohort of refugee children in a regional community in Australia." BMJ Open **6**(8).

McMichael, C., et al. (2015). "Studying Refugee Settlement through Longitudinal Research: Methodological and Ethical Insights from the Good Starts Study." Journal of Refugee Studies **28**(2): 238-257.

Gibbs, L., et al. (2014). "An exploratory trial implementing a community-based child oral health promotion intervention for Australian families from refugee and migrant backgrounds: a protocol paper for Teeth Tales." BMJ Open **4**(3): e004260.

*Theme: Advocacy*

Dechent, S., et al. (2019). "Asylum Seeker Children in Nauru: Australia's International Human Rights Obligations and Operational Realities." International Journal of Refugee Law **31**(1): 83-131.

Essex, R. and D. Isaacs (2018). "The Ethics of Discharging Asylum Seekers to Harm: A Case From Australia." Journal of Bioethical Inquiry **15**(1): 39-44.

Nardone, M. and I. Correa-Velez (2016). "Unpredictability, Invisibility and Vulnerability: Unaccompanied Asylum-Seeking Minors' Journeys to Australia." Journal of Refugee Studies **29**(3): 295-314.

Woodhead, M. (2016). "Australian doctor challenges government over child detention "torture"." BMJ **352**: i550.

Thomas, L. (2016). "The Lost Souls: How Australia's Asylum Seeker Policy is Damaging Children." Aust Nurs Midwifery J **23**(9): 16-21.

Mares, S. (2016). "Fifteen years of detaining children who seek asylum in Australia - evidence and consequences." Australas Psychiatry **24**(1): 11-14.

Rowcliffe, C., et al. (2016). "The impact of detention on children and adolescents." J Paediatr Child Health **52**(9): 912-913.

Zwi, K. and S. Mares (2015). "Stories from unaccompanied children in immigration detention: A composite account." J Paediatr Child Health **51**(7): 658-662.

Paxton, G., et al. (2015). "Perspective: 'The forgotten children: national inquiry into children in immigration detention (2014)'." J Paediatr Child Health **51**(4): 365-368.

Isaacs, D. (2015). "Nauru and detention of children." J Paediatr Child Health **51**(4): 353-354.

Corbett, E. J., et al. (2014). "Australia's treatment of refugee and asylum seeker children: the views of Australian paediatricians." Med J Aust **201**(7): 393-398.

Zwi, K. and S. Mares (2014). "Commentary: reducing further harm to asylum-seeking children. The global human rights context." Int J Epidemiol **43**(1): 104-106.

Zwi, K. and G. Chaney (2013). "Refugee children: rights and wrongs." J Paediatr Child Health **49**(2): 87-93.

Dudley, M., et al. (2012). "Children and young people in immigration detention." Current Opinion in Psychiatry **25**(4): 285-292.

Cameron, G., et al. (2011). "Young Refugees in Australia: Perspectives From Policy, Practice and Research." Children Australia **36**(2): 46-55.

Jureidini, J. and J. Burnside (2011). "Children in immigration detention: a case of reckless mistreatment." Aust N Z J Public Health **35**(4): 304-306.

*Theme: Health literacy*

Joseph, J., et al. (2019). ""Fitting-in Australia" as nurturers: Meta-synthesis on infant feeding experiences among immigrant women." Women and Birth **32**(6): 533-542.

Joseph, J., et al. (2019). "From Liminality to Vitality: Infant Feeding Beliefs Among Refugee Mothers From Vietnam and Myanmar." Qual Health Res: 1049732318825147.

Fernandes, S., et al. (2015). "What makes people sick? Burmese refugee children's perceptions of health and illness." Health Promot Int **30**(1): 151-161.

Watts, M., et al. (2015). "Factors Influencing Contraception Awareness and Use: The Experiences of Young African Australian mothers." Journal of Refugee Studies **28**(3): 368-387.

Wilson, A. and A. Renzaho (2015). "Intergenerational differences in acculturation experiences, food beliefs and perceived health risks among refugees from the Horn of Africa in Melbourne, Australia." Public Health Nutrition **18**(1): 176-188.

Ngum Chi Watts, M. C., et al. (2014). "Contraception knowledge and attitudes: truths and myths among African Australian teenage mothers in Greater Melbourne, Australia." J Clin Nurs **23**(15-16): 2131-2141.

De Anstiss, H. and T. Ziaian (2010). "Mental health help-seeking and refugee adolescents: Qualitative findings from a mixed-methods investigation." Australian Psychologist **45**(1): 29-37.
